# Clinical utility of repurposing a short course of hepatitis C drugs for COVID19. A randomized controlled study

**DOI:** 10.1101/2022.07.18.22277477

**Authors:** Mostafa Yakoot, Basem Eysa, Essam Gouda, Sherine A Helmy, Mahmoud R Elsayed, Ola Elrouby, Amal Mohamed, Ehab Reyad, Mohamed M Fawzi, Safaa Elmandouh, Wessam Abdellatif, Ahmed A Zidan, Abd-Elmoniem Adel, Mohamed Hassany

**Affiliations:** Green Clinic and Research Centre, Alexandria University, Egypt; The National Hepatology and Tropical Medicine Research Institute (NHTMRI), Cairo, Egypt; Chest Diseases Department, Alexandria Faculty of Medicine, Alexandria, Egypt; Pharco Corporate, Alexandria, Egypt

**Keywords:** COVID-19, Drug therapy, Sofosbuvir, Daclatasvir, Randomized controlled study

## Abstract

**BACKGROUND:** Preliminary data suggests a potential therapeutic benefit for the hepatitis C drugs, sofosbuvir (SOF) and daclatasvir (DCV) for the treatment of COVID-19. We aim to evaluate efficacy of a short course of dual sofosbuvir/daclatasvir in patients with COVID-19.

**METHODS:** Eighty-nine consecutive eligible patients were randomly assigned to two treatment groups. The experimental group was treated with the standard of care (SOC) therapy in addition to one 400 mg tablet sofosbuvir and one 60 mg daclatasvir daily for 10 days; while the control group was treated with the SOC therapy alone. Baseline clinical data was measured and followed up for 21 days. Data was compared between the two treatment groups.

**RESULTS:** The proportion of cumulative clinical recovery in the experimental group at day 21 was numerically greater than the control group (40/44 (91%; 95%CI: 78.8-96.4%) versus 35/45 (77.8%; CI 63.7-87.5%)). The Hazard Ratio (HR) for time to clinical recovery adjusted for baseline severity, using a Cox-regression model was statistically significant: HR: 1.59 (95%CI: 1.001-2.5). Concordantly, the experimental group also showed trends for greater improvement in the mean 8-points ordinal scale score, the severity of lung lesions score and the case fatality rate (4.5% versus 11.1%). No serious or severe adverse events were reported in both groups.

**CONCLUSION:** This study supports potential benefit and safety of sofosbuvir combined with daclatasvir when given early in the treatment of COVID-19. We hope to encourage further large sized, multinational studies to confirm the results.

**HIGHLIGHTS:**

- Preliminary data suggests a potential therapeutic benefit for the hepatitis C drugs, sofosbuvir (SOF) and daclatasvir (DCV) for the treatment of COVID-19.
- Eighty-nine COVID-19 patients were randomly assigned to either treatment with SOC plus a short course of combined SOF/DCV therapy or SOC therapy alone.
- The Hazard Ratio (HR) for time to clinical recovery adjusted for baseline severity showed statistical significance: HR: 1.59 (95%CI: 1.001-2.5). Concordantly, all other efficacy endpoints showed numerical trends for greater improvement in the experimental group including the case fatality rate (4.5% versus 11.1%). No serious or severe adverse events were reported in both groups.
- SOF/DCV therapy might be beneficial when given early in the treatment of COVID-19.

## INTRODUCTION

Coronavirus disease 2019 (COVID-19), caused by a novel human coronavirus (SARS-CoV-2) is a public health emergency of international concern. There is an urgent need for readily available effective pharmaceutical treatments to help save lives of severely infected patients and to reduce the infectiousness of early and mild cases to others by reducing viral loads in the respiratory secretions of patients.[1] SARS-CoV-2 is an enveloped positive-sense, single strand RNA virus that shares similar replication mechanisms to other single-stranded RNA viruses such as the hepatitis C virus (HCV). [2] Given the success with HCV, it was thought that the virus RNA-dependent RNA polymerase (RdRp) and the replicase enzyme systems could be among the important targets for therapy for SARS-CoV-2. [3] Conventional drug development takes time and so repurposing existing pharmaceuticals offers a pragmatic alternative in responding quickly and efficiently to the COVID-19 pandemic. Sofosbuvir is an example of an existing nucleotide analogue that acts through inhibition of the RNA-dependent RNA polymerase of the hepatitis C virus by competing with natural ribonucleotides. [4] Sofosbuvir has shown efficacy in the treatment of chronic HCV combined with many other direct acting antiviral drugs such as ledipasvir and daclatasvir. [5] It was suggested that acting through the same mechanisms, sofosbuvir combined with ledipasvir, daclatasvir or velpatasvir might be able to also inhibit SARS-CoV-2. This has been supported by data from a molecular docking experiment that identified tight binding of sofosbuvir to the SARS-CoV-2 RdRp [6]. In similar *in silico* studies, daclatasvir was also shown to bind to SARS-CoV-2. [7]

Furthermore, in-vitro studies showed that sofosbuvir inhibited the production of infectious SARS-CoV-2 virus particles in the hepatoma cell line (HuH-7) and in type II pneumocytes (Calu-3), with EC_50_ values of 5.1 and 7.3 μM respectively [8]. Sofosbuvir terminated RNA was shown to display resistance to excision by the SARS-CoV-2 exonuclease-based proofreader. The addition of DCV to SOF was shown to augment this resistance and to increase the potency of SOF 10-folds causing reduction of EC50 to 0.7 μM in Calu-3 cell line. [8-10] Sofosbuvir was also shown to protect neural cells from SARS-CoV-2-induced cell death in human brain organoids [11]. Daclatasvir also showed inhibitory activity against SARS-CoV-2 with EC_50_ values of 1.1 μM, 0.8 μM and 0.6 μM in Calu-3, Vero E6 and Huh-7 cell lines respectively [8]. Daclatasvir was also found to prevent the enhancement of IL-6 and TNF-α levels in human monocytes. [8]. At standard human dosing, EC_50_ estimates for daclatasvir are within pharmacokinetic exposures. Results from small clinical studies in Iran demonstrated the efficacy of sofosbuvir/daclatasvir combination therapy in the treatment of severe cases of COVID-19 with reduction in mortality [12-14].

### STUDY OBJECTIVES

Evaluate the efficiency and safety of sofosbuvir/daclatasvir in addition to the standard of care therapy (SOC) versus the standard of care alone in patients with 2019-nCoV infection, initially hospitalized to a non-intensive care setting.

## METHODS

### STUDY DESIGN & SETTING

This is a parallel 2-arm, open-label, randomized controlled clinical study. The study has been conducted in the isolation hospital of the National Hepatology and Tropical Medicine Research Institute (NHTMRI) in Cairo, under the guidance, monitoring and supervision of the clinical investigator team and the ministry of health committee for the control of COVID-19.

### PATIENTS

Eighty-nine consecutive patients presenting to one of the isolation hospitals in Cairo (NHTMRI), with suspected or confirmed COVID-19 infection were included in the study and randomized after fulfilling all the eligibility criteria and signing the informed consent form. The study protocol was reviewed and approved by the Research Ethics Committee of Faculty of Medicine, Alexandria University (IRB00007555) and the Central Egyptian Ministry of Health and People Research Ethics Committee and registered in the German clinical trial database repository (DRKS00022203) before the study initiation.

Participants included in the study were between 18 and 75 years of age and presented with laboratory-confirmed COVID-19 infection as determined by polymerase chain reaction (PCR) assay in any specimen collected within 72 hours prior to randomization. Inclusion criteria also included being symptomatic at screening, presenting with clinical manifestations under one of three severity categories: Mild, Moderate or Severe disease. Mild was defined as having mild symptoms with no picture of pneumonia in Computerized Tomography scan (CT-scan). Moderate was defined as having moderate fever and respiratory symptoms plus evidence of pneumonic lesions in CT-scan. The severe disease (but Not Critical) included patients with laboratory confirmed COVID-19, presenting at baseline by any of the following criteria: respiratory distress and respiratory rate (RR)≥30 times/min; Finger oxygen saturation ≤93% in rest state; Arterial partial pressure of oxygen / concentration of fractional inspired oxygen (PaO2/FiO2) ≤300mmHg and > 200mmHg under oxygen inhalation not necessitating ICU admission or mechanical ventilation.

The main exclusion criteria were refusal or inability to sign the informed consent form; pneumonia due to other etiology; patients with acute respiratory distress syndrome requiring invasive mechanical ventilation at screening; late presentation of patient after 10 days from the appearance of symptoms; severe concomitant illness that affects survival or course of the disease including uncontrolled malignancy, HIV, active bleeding, shock/or multiple organ failure at screening. Further exclusion criteria included pregnancy or lactating females; hypersensitivity or contraindication to any of the experimental drugs used in the study (Prolonged QT syndrome, G6PD deficiency, psoriasis, retinal damage or others); decompensated liver cirrhosis or abnormal liver enzyme tests above three times the upper limit values (alanine aminotransferase – (ALT) and aspartate aminotransferase (AST); renal dysfunction (estimated glomerular filtration rate [eGFR] <30 mL/min/1.73m^**2**^).

### RANDOMIZATION AND STUDY MEDICATIONS

All patients who fulfilled the eligibility criteria at screening with confirmed positive PCR results were included in the study and admitted to the hospital.

Patients were stratified according to baseline severity into three groups and randomized by a computer generated block-randomization technique (1:1) to either one of two treatment groups: **Group 1 (experimental)** received the standard of care (as per the Egyptian Ministry of Health (MOH) protocol effective at the time of recruitment [15]) together with a daily dose of one Gratisovir (sofosbuvir) 400 mg tablet combined with one Daktavira (daclatasvir) 60 mg tablet for 10 days, (both are generic products produced by Pharco Corporate, Alexandria, Egypt). **Group 2 (control /Active Comparator)**, received the standard of care therapy (according to the Egyptian MOH protocol) for 10 days. The randomization was conducted centrally by a statistician not involved in patient care and the random allocation sequence was concealed by opaque sealed envelopes.

The standard of care therapy (SOC), according to the Egyptian MOH Protocol, which was given to all patients in both groups included: Plaquenil (hydroxychloroquine) given in a dose of 4 tablets of 200 mg on day 1 followed by 2 tablets daily from day 2 through day 5. Azithromycin was given in a daily dose of 500 mg for 5 days. Full nutritional support with balanced diet and multivitamin and zinc supplements with vitamin C and D were offered to all patients. Other drugs were given according to each patient’s clinical condition and included: analgesic-antipyretic (acetaminophen 500 mg as needed); cough mixtures: either a cough suppressant with low dose dextromethorphan (for irritating dry cough) or a mixture of a mucolytic (ambroxol) combined with a small theophylline dose (Farcosolvin) aiming to provide enhancing effect on muco-ciliary escalation and surfactant release from type-2 alveolar cells.

### PROCEDURES

All randomized patients were subjected to full medical history taking clinical examination, routine laboratory tests and appropriate imaging studies to record the baseline data. All patients were monitored daily for any change in clinical manifestations, including symptom scoring for severity, vital signs, temperature, pulse, blood pressure, EKG, respiratory rate and oxygen saturation by pulse oximeter. This allowed for the measurement of time to clinical recovery and the progress in clinical status using the 8-points ordinal scale over time during the study period. The 8-point-ordinal-scale was measured daily and scored as follows: 1) Death; 2) Hospitalized, on invasive ventilation or ECMO; 3) Hospitalization on NIV or HFO2; 4) Hospitalized with oxygen supplement; 5) Hospitalized, not requiring oxygen but need medical care; 6) Hospitalized, not requiring oxygen or immediate medical care; 7) Not hospitalized but with limitation on activities and/or requiring home oxygen; 8) Not hospitalized with normal activity. Routine laboratory tests including complete blood count, CRP, serum ferritin, D-Dimer, Lactate dehydrogenase, Troponins, liver and kidney functions, were repeated every other day. Nasal/throat swabs were taken on days 0, 3, 5, 10, 14 and 21. Imaging studies by chest computerized tomography scan (CT scan) and ultrasonography for chest and abdomen were taken throughout the course of the study on a twice weekly basis. Any emerging medical conditions or adverse events encountered were dealt with on a case-by-case basis with appropriate laboratory, imaging studies or therapeutic interventions. [16]

Treatment-emergent adverse event (TEAE) was defined as any adverse event reported by patients or observed by investigators during the study visits or any deviation from a baseline normal laboratory test, occurring after administration of the first dose of study drugs until 30 days after the last dose. All TEAEs were reported using the Medical Dictionary for Regulatory Activities version 22 (MedDRA v.22) lowest level and preferred terms. TEAEs deemed to have certain, probable or possible causality according to Uppsala Monitoring Center (UMC) causality categorization were considered in analysis for incidence rate in an intention-to-treat basis and graded according to seriousness (serious/non-serious) and severity using the Common Terminology Criteria for Adverse Events v3.0.

### OUTCOME MEASURES

The main primary endpoints of this trial were proportions-of and time-to-clinical recovery within the 21day time-frame following enrolment. Clinical recovery was defined as normalization of fever (≤37.2°C), respiratory rate (≤24 breaths/min) and oxygen saturation (≥ 94% on room air), sustained for at least 24 hours with improvement of at least 2 points at the WHO-8-point-ordinal-scale (described above in procedure section). Other endpoints included the mean change in clinical status using the 8-point-ordinal-scale; the proportion of patients achieving sustained viral negativity assessed twice 48 hours apart (by reverse transcription-PCR) during hospital stay; the effect of treatment on the progression/improvement of lung lesions using repeated-measures-factorial-ANOVA test with adjustments for baseline disease severity, diabetes, CRP; [16, 17] Ferritin and D-Dimer as covariates; and proportion of patients who deteriorated and required escalation of treatment to mechanical ventilation or died during the study follow up period. [15-21]

### STATISTICAL METHODS

Group sample sizes of 44 in group one and 44 in group two achieve 81% power to detect a difference between the group proportions of 0.24. The proportion in group one (the treatment group) is assumed to be 0.66 under the null hypothesis and 0.90 under the alternative hypothesis. The test statistic used is z test for proportions with boot strapping. The significance level of the test was targeted at 0.05. The primary indicators collected in this study are described with descriptive statistical methods. Quantitative variables are described as mean, standard deviation, median and interquartile range. Qualitative indicators are described by frequency, percentage and the proportions with 95% confidence interval (CI) using Wilson Score Interval Method. The statistical significance level is 0.05 by two-sided test (one-sided 0.025) and the 95% confidence intervals were provided for the estimation of inter-group variance parameters. Data has been analyzed using the computer software SPSS version 21. Scale variables were compared by repeated measures factorial one-way analysis of variance. Exact test with boot strapping was used for analysis of some of the categorical variables.

A Kaplan –Meier curve was used to report the probability of progression over time (time to clinical recovery). The hazard function for time to recovery in both treatment groups were compared using Cox regression with adjustment for baseline disease severity. All statistical tests were based on the intention-to-treat population analysis.

## RESULTS

Between June and September 2020, eighty-nine patients participated in the study, they were all randomized and received their assigned medications (Figure 1. Patient Flowchart), forty-four patients were randomized to sofosbuvir/daclatasvir add-on to the standard of care therapy (experimental group) and forty-five patients were randomized to the standard of care therapy alone (control group) “as fully described in methods section”. All patients were adults and their age (median (IQR)) was (48 (31-59)) years. Baseline laboratory parameters were balanced across arms, which included comorbidities and baseline severity categories (Table 1).

**Table 1:** Baseline characteristics.

| PARAMETERS | EXPERIMENTAL<br>(n=44) | CONTROL<br>(n=45) |
| --- | --- | --- |
| **Baseline demographics** |  |  |
| Age, median (IQR) | 48 (34-59) | 50 (31-60) |
| Male/female, n (%) | 18/26 (41% / 59%) | 20/25 (45% / 55%) |
| Severity of disease at baseline, n (%) |  |  |
| Mild | 6 (14%) | 6 (13%) |
| Moderate | 30 (68%) | 31 (69%) |
| Severe | 8 (18%) | 8 (18%) |
| Comorbidities, n (%) |  |  |
| Asthma | 0 (0%) | 1 (3%) |
| Diabetes | 9 (22%) | 8 (21%) |
| Chronic kidney disease | 0 (0%) | 2 (4%) |
| Hypertension | 11 (25%) | 12 (27%) |
| Heart disease | 2 (4%) | 6 (13%) |
| Smoking | 12 (29%) | 7 (18%) |
| Vital signs at baseline, median (IQR) |  |  |
| O<sub>2</sub> saturation (%) | 96 (94-97) | 96 (94-98) |
| Temperature (°C) | 37 (36.8-37.8) | 37 (36.9-37.2) |
| Respiratory rate (Breaths/min) | 19 (18-24) | 20 (18-25) |
| Pulse (Beats/min) | 91 (82-100) | 90 (85-98) |
| Laboratory findings on admission, median (IQR) |  |  |
| Lymphocytes | 1.9 (1.3-2.4) | 2.1 (1.3-2.6) |
| D-Dimer (mg/L) | 0.46 (0.31-0.59) | 0.56 (0.33-1.05) |
| C-Reactive protein (mg/dL) | 22.2 (4.8-66.4) | 23.4 (4.0-70.1) |

**Figure 1:**
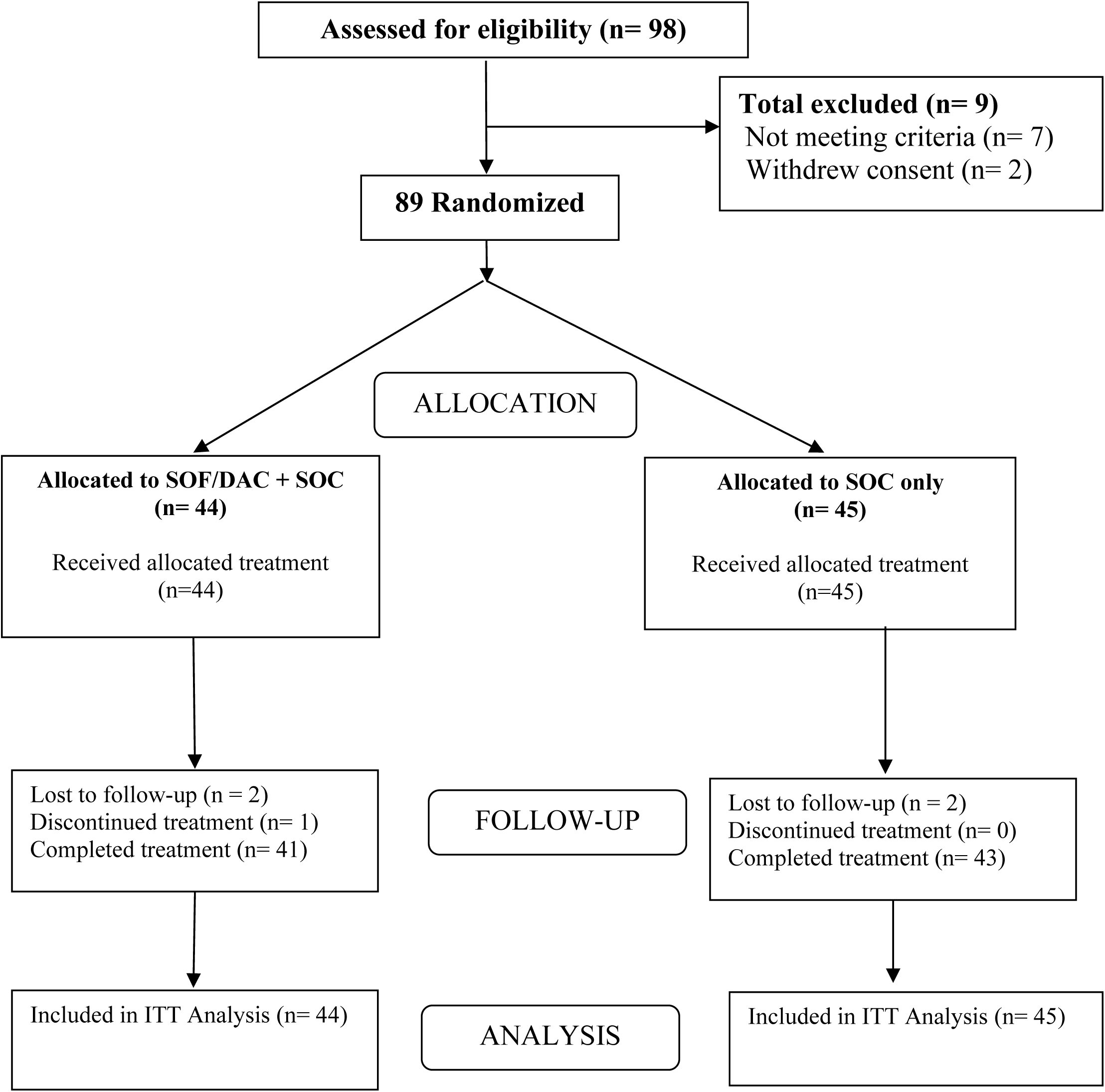
Patient Flowchart.

During the course of the study, all patients were subjected to daily monitoring for their vital signs, and clinical data for accurately measuring the time-to-clinical recovery, the 8-point-ordinal-scale scores. Other laboratory and imaging studies were performed at the scheduled times according to the protocol.

After 21 days from the start of therapy, forty Patients (90.9%; 95% confidence interval (CI): 78.8-96.4%) in the experimental group and thirty-five patients (77.8%; 95% CI: 63.7-87.5%) in the control group fulfilled the criteria of clinical recovery (afebrile, no tachypnea, O2 saturation ≥ 94% on room air, improved or stable chest lesions in multi-slice CT-scan for chest) (Table 2). Seventeen out of eighteen males (94.4%) in the experimental group achieved clinical recovery at day 21; while twenty-three out of twenty-six (88.5%) females in the experimental group achieved clinical recovery at day 21 (Table 2). Sex appeared to have no significant effect on both the clinical recovery and fatality at day 21 (p > 0.05) while age in years as a scale quantitative variable showed negative correlation with the clinical recovery and positive correlation with fatality at day 21 by logistic regression (p < 0.05) as shown in table 3.

**Table 2:** Clinical Outcomes.

| **Clinical Outcome** | **SOF/DCV**<br>**n=44** | **Control**<br>**n=45** | **P value**<br>**(2 sided)** |
| --- | --- | --- | --- |
| Clinical Recovery at day 21 (Total), n (%) | 40 (90.9%) | 35 (77.8%) | 0.14 <sup>a</sup> |
| Clinical Recovery, day 21 (male), k/n (% within males) | 17/18 (94.4%) | 15/20 (75%) | 0.18 <sup>a</sup> |
| Clinical Recovery, day 21 (Female), k/n (% within females) | 23/26 (88.5%) | 20/25 (80%) | 0.46 <sup>a</sup> |
| Time to clinical recovery, mean (95% CI) | 7 (5.5-8.6) | 9.4 (7.4-11.4) | 0.059 |
| Time to clinical recovery, HR (95% CI)<br>(adjusted for baseline severity) | 1.59 (95% CI: 1.001-2.5) |  | 0.049* |
| **Case Fatality, k/n (%)** |  |  |  |
| Overall (Total patients) | 2/44 (4.5%) | 5/45 (11%) | 0.43 |
| Mild baseline severity | 0/6 (0%) | 0/6 (0%) |  |
| Moderate baseline severity | 1/30 (3.3%) | 3/31 (9.7%) |  |
| Severe baseline severity | 1/8 (12.5%) | 2/8 (25%) |  |
| <sup>a</sup> non-significant using 2 sided exact test<br>P value for time to recovery analysis calculated using log-rank test.<br>P value for HR-time to clinical recovery adjusted for baseline severity calculated by Cox-regression test.<br>*(Statistically Significant).<br>P value for categorical outcomes calculated using Fisher's exact test. |  |  |  |

**Table 3:** Regressions of Clinical Recovery and Fatality at day 21 on age, sex and type of treatment.

| Independent variables | B (Slopes) |  | Sig. |  | Odds Ratios |  | 95% CI for EXP(B) |  |
| --- | --- | --- | --- | --- | --- | --- | --- | --- |
|  | Recovery | Fatality | Recovery | Fatality | Recovery | Fatality | Recovery | Fatality |
| Treatment | 1.206 | -1.097 | 0.076 | 0.23 | 3.338 | 0.33 | 0.88 - 12.62 | 0.056 - 2 |
| Sex (male/female) | 0.236 | -0.325 | 0.717 | 0.71 | 1.267 | 0.72 | 0.35 - 4.54 | 0.131- 3.98 |
| Age in years (scale) | - 0.079 | 0.103 | 0.006* | 0.023* | 0.924 | 1.1 | 0.87 - 0.98 | 1.01 - 1.21 |
| Constant | 5.182 | -7.418 | 0.001 | 0.005 | 177.97 | 0.001 |  |  |

a. Dependent variables are Clinical Recovery and Fatality at day 21; Independent variable(s) entered: Treatment, Sex, Age. \* (Statistically Significant).

In order to further evaluate the effect of treatment on clinical recovery, we tested the time to clinical recovery in both groups, using the Kaplan Meier, Log-rank, and Cox-regression tests. The experimental group was found to have numerically faster mean time to clinical recovery than the control group (7 versus 9.42 days). The difference was close to margin of statistical significance by unadjusted log rank test, p =0.059 (Table 2), but the Hazard Ratio adjusted for baseline severity, estimated by Cox-regression, was statistically significant, hazard ratio 1.59 (95% CI: 1.001-2.5), p=0.049 (Figure 2).

**Figure 2:**
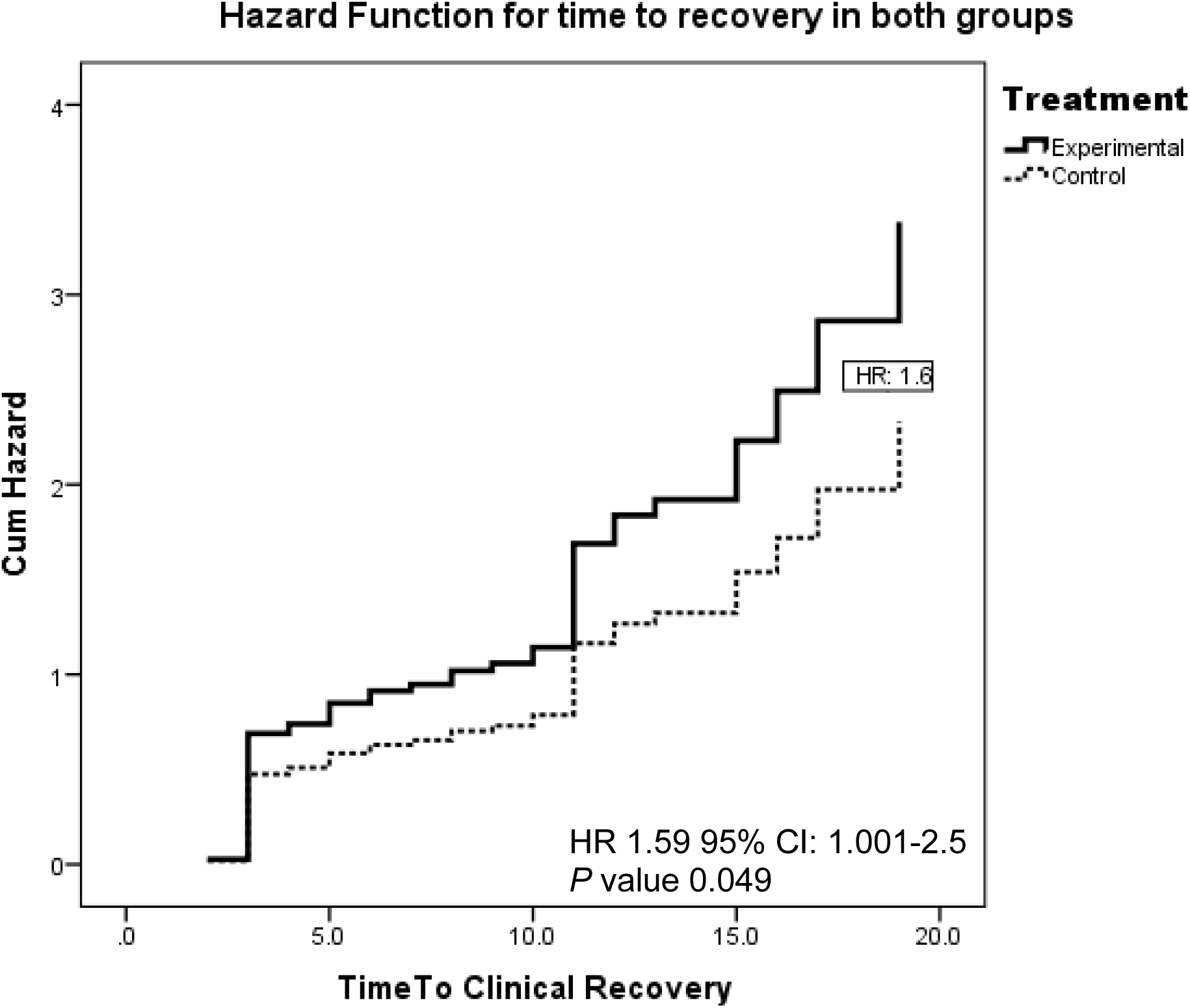
Hazard Function for time to clinical recovery in both treatment groups.

We also compared the change in mean of the 8-point-ordinal-scale scores between the two treatment groups at 4 time points during the course of the study: at day 0 (baseline), day 5, 14 and 21 (end of follow up period), using repeated measures, factorial analysis of variance, (ANOVA test) adjusted for important baseline severity factors (baseline: severity, O2 saturation, CRP, D-Dimer). The mean 8-point-ordinal-scale score showed higher numerical improvement in the experimental group (Figure 4), with the curves starting to diverge from day 5, but this did not reach statistical significance at the study sample size in the intent-to-treat (ITT) analysis, p=0.31

**Figure 3:**
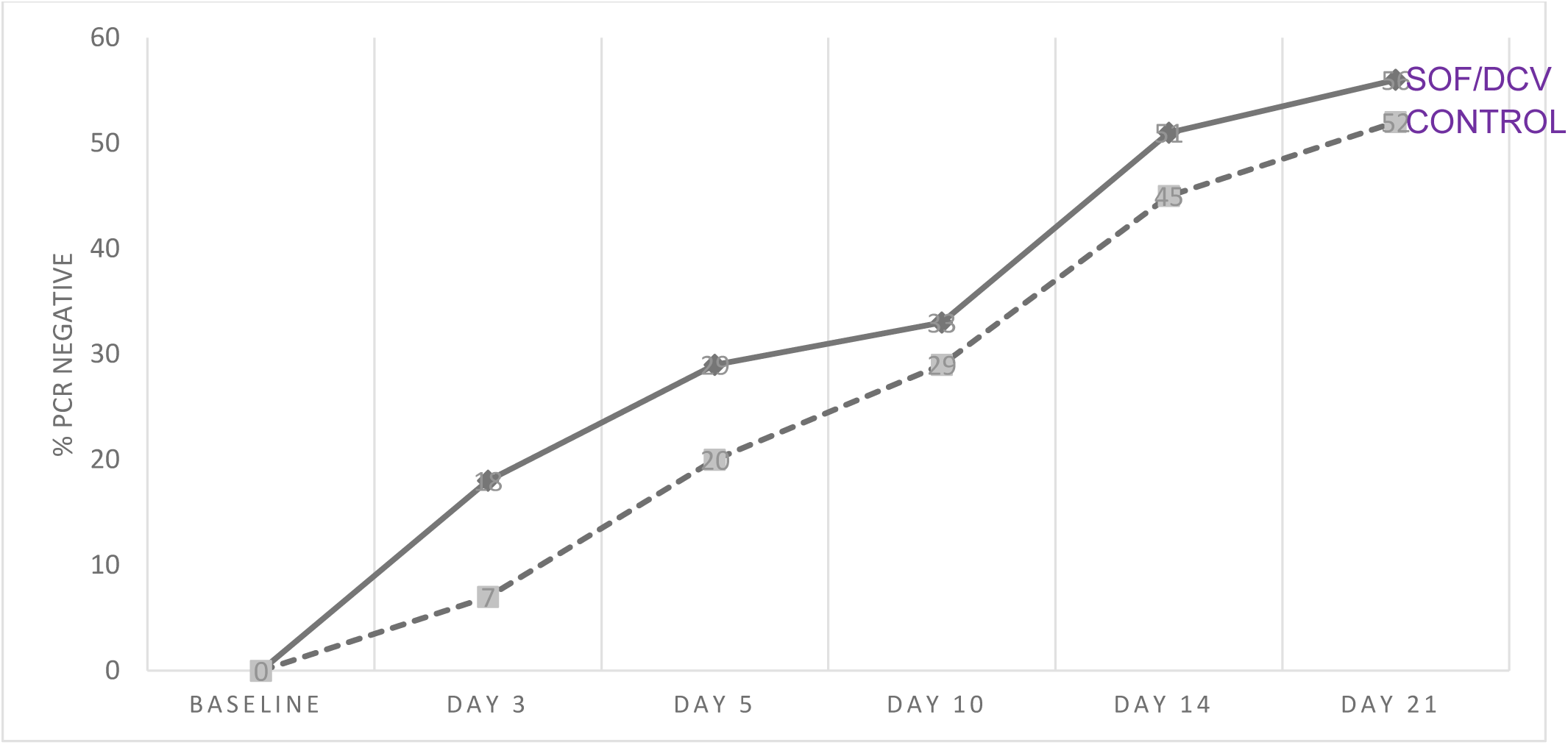
The percentage of viral negativity over time using an intent-to-treat approach (missing data was classified as a failure).

**Figure 4:**
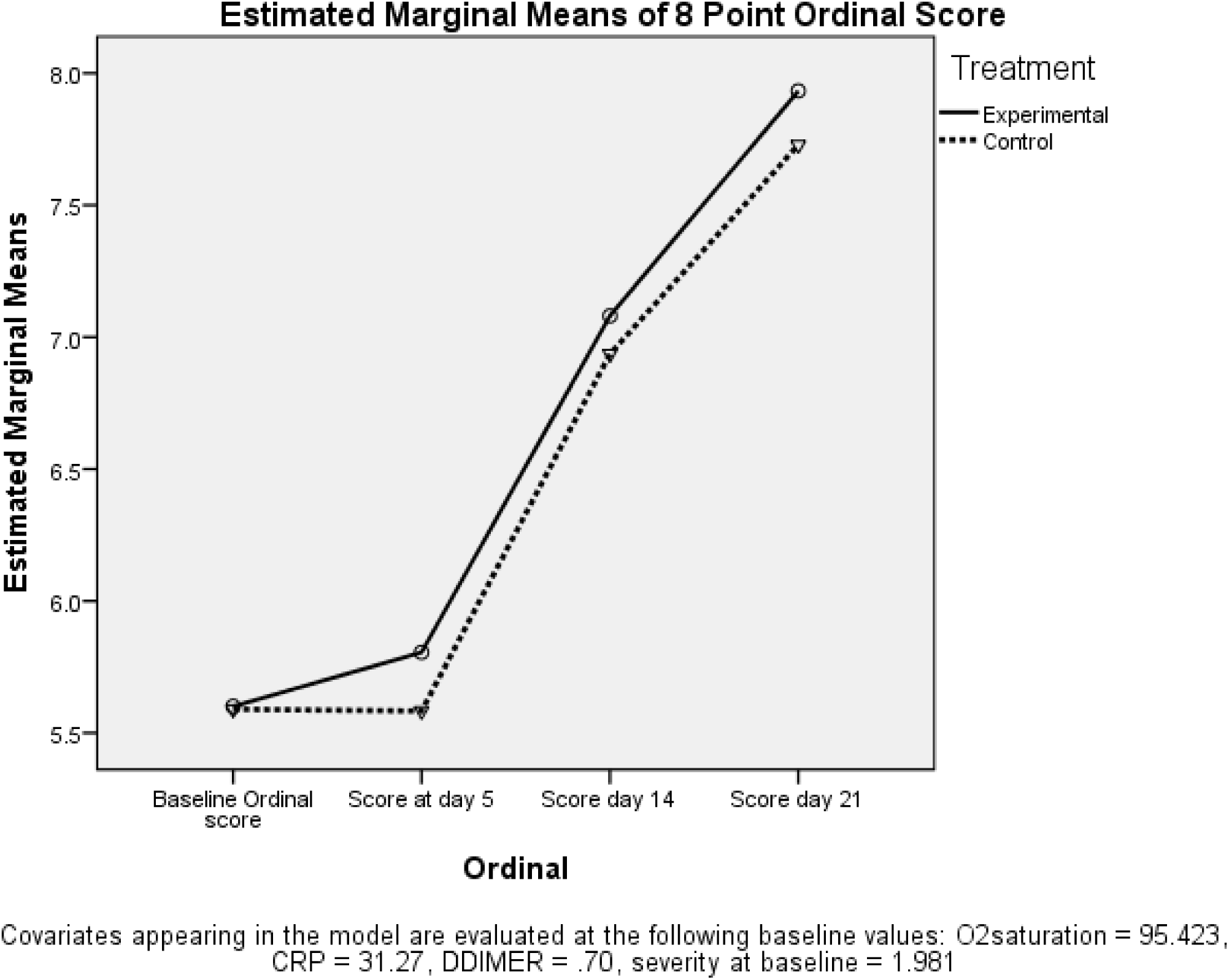
Mean values of the 8-point ordinal scores in both treatment groups.

Multi-slice computerized tomography scans (CT-scans) were performed for all patients at baseline, and repeated according to the protocol follow-up schedule. Lung lesions were evaluated for size, extension and number, and were given scores from 0% (normal lung) up in severity to 100%.

Repeated-measures-factorial-ANOVA test was done to compare the effect of treatment on the progression/improvement of lung lesions; with adjustments for baseline disease severity, history of diabetes, CRP, Ferritin and D-Dimer as covariates. Figure 5 depicts the mean severity scores of lung lesions in both treatment groups at baseline, day 10 and day 21. The experimental group showed tendency for improvement in lung lesions with greater reduction in the mean severity scores (numerically), though the difference was not statistically significant (p= 0.65).

**Figure 5:**
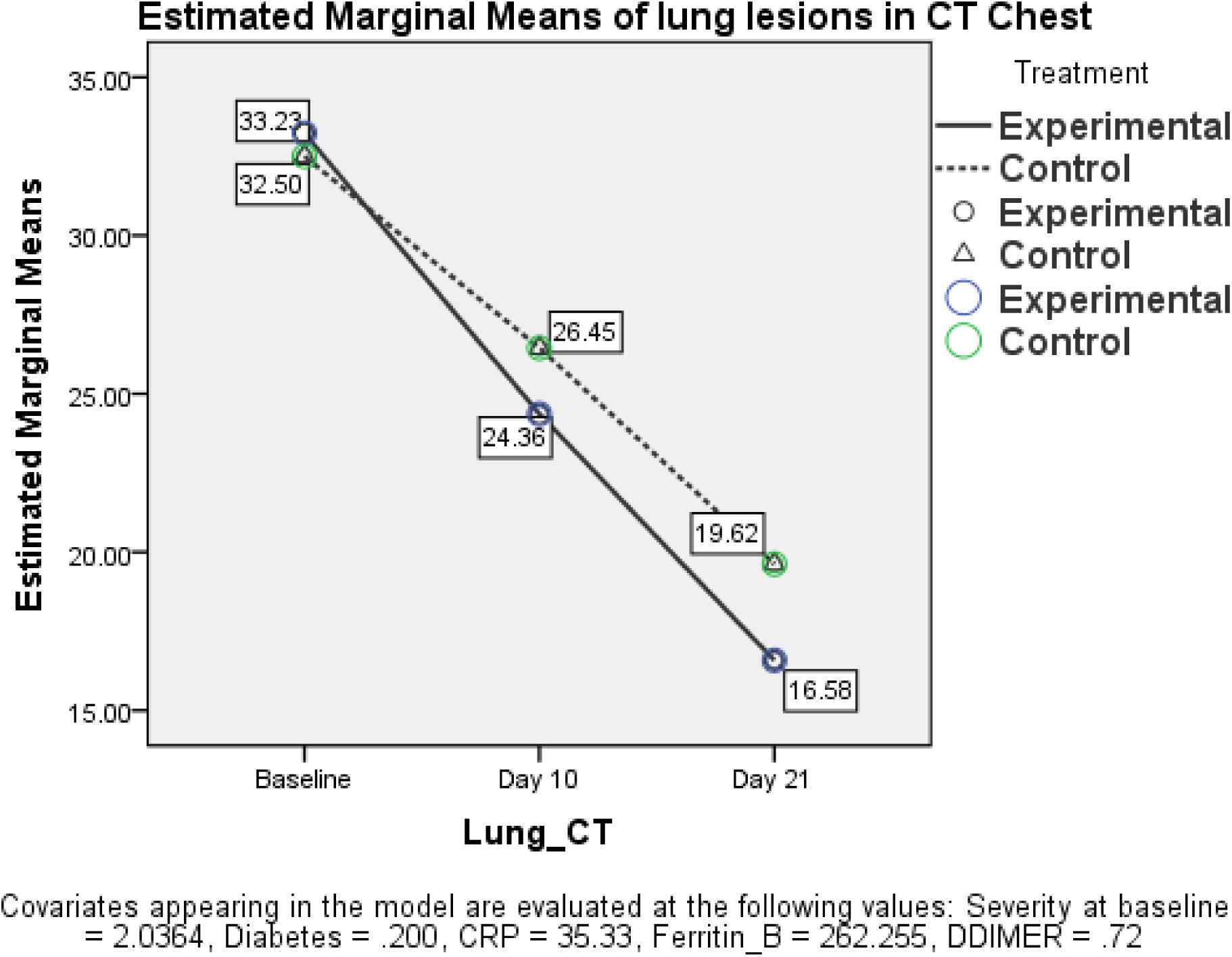
Mean severity scores of lung lesions in both groups at three time points.

Figure 3 shows the percentage of individuals who became PCR negative for nCov2 virus during the follow-up period. This figure uses an-intent-to-treat approach; whereas missing values were classified as failures.

During the course of the study, two patients in the experimental group (4.5%; 95% CI: 1.13 - 15.1%) and five patients in the control group, (11.1%; 95%CI: 4.8 - 23.5%) experienced severe deterioration in their condition, resulting in their admission to ICU, where they were put on invasive mechanical ventilation. All seven of these patients died in ICU (Table 2), and were reported by the attending physicians to have died due to complications of the underlying medical condition rather than the study medications.

No serious or severe adverse events (grade three or above) were reported in both treatment groups. Few non-serious adverse events were reported in both groups. Five patients reported palpitations and transient tachycardia, (three patients in the control group and two patients in the experimental group). Otherwise, transient mild complaints of nausea, abdominal pain were recorded in two patients in each group, headache by one patient in the experimental group, elevated liver transaminases by one patient in the control group and skin rash reported by one patient in the experimental group.

## DISCUSSION

In this randomized controlled trial on mild to severe hospitalized COVID-19 patients, we showed that the combination of sofosbuvir/daclatasvir might shorten the time to clinical recovery when adjusting for baseline severity compared to standard care alone (Hazard Ratio (HR) adjusted for baseline severity, estimated by Cox-regression, was statistically significant, hazard ratio 1.59 (95% CI: 1.001-2.5), p=0.049 (Figure 2). This signifies that, at any time point during the study, the clinical recovery in the experimental group (SOF/DCV group) has about 1.6 times greater probability than the control group. Sex (male or female) appeared to have no significant effect on both the clinical recovery and fatality at day 21, while age showed significantly negative correlation with the clinical recovery (the younger the age the better the chance for recovery) and positive correlation with fatality (table 2 and 3). Other parameters, such as the case fatality rates, the mean change in the 8-points-ordinal-scale scores, and lung lesions severity scores also showed numerical trends for better improvement in experimental group. Though these trends did not reach statistical significance at the studied sample size, all were concordant with the statistically significant HR (hazard-function for time-to-clinical-recovery). Being all concordant in one direction (in favor of experimental group) could be considered at least a signal against the mere chance as a cause for better outcome in experimental group. This, in our viewpoint, could strengthen the evidence for the potential benefits of SOF/DCV as add-on treatment for COVID-19 patients.[22]

The highly variable outcome of COVID-19, with its considerable proportion of spontaneous recovery and low unpredictable fatality rate in untreated mild to moderate cases, could make the effect of any intervention on hard endpoints such as case fatality rate difficult to prove in small-sized clinical trials. Even in large-sized, randomized controlled studies, it was practically difficult to prove the reduction in mortality beyond the standard of care (control group) with the conventional statistically significant tests. In our opinion, the time to initiate antiviral drug therapy during the course of the disease is the major determinant factor for antiviral efficacy. Starting antiviral therapy during the early phase of rapid viral replication (first week of symptoms) is the ideal time to test antiviral efficacy, while most clinical studies include patients during the second or third phase of the disease when viral replication becomes no more important, as the patients become more under attack from their own immune systems. The case fatality rate in the experimental group in our study was numerically lowered by more than 50% (4.5% in experimental group versus 11.1% in the control group), yet as in most other antiviral studies for COVID-19, it did not reach statistical significance.[22]

From basic biostatistics understanding, to detect a statistically significant difference in case fatality rate between groups included in the early phase of infection or with mild disease severity, it requires very large sample size (because the fatality rate in such population is already small), hence we often see non-significant trends.

On the other hand, in studies on advanced or critical cases, antiviral drugs become of less biological plausibility and not lifesaving. Other heterogeneous uncontrollable factors such as the behavior of the genetically-determined host-inflammatory-response, the appropriate use of anti-inflammatory drugs, O2, ventilators, good nursing and other critical care interventions become more important independent lifesaving variables at late stages.[22]

A number of antivirals have been assessed against COVID-19, such as favipiravir [23,24], lopinavir/ritonavir [24,25], hydroxychloroquine [24, 26] and remdesivir [24]. However, no conclusive evidence of mortality benefit was shown even in large clinical trials. Results from the WHO SOLIDARITY trial showed that remdesivir, hydroxychloroquine and lopinavir/ritonavir show no benefit in reducing mortality or duration of hospitalization. [25] Furthermore, in the UK RECOVERY trial, no clinical benefit was associated with hydroxychloroquine or lopinavir/ritonavir and randomizations to these arms were stopped. [26, 27]. The corticosteroid dexamethasone has shown survival benefit in severe patients receiving oxygen. [28] Thus, a strong necessity to identify effective antiviral treatments remains.

This study together with promising results from three clinical trials conducted in Iran [12-14], could provide preliminary support for the potential benefit of sofosbuvir combined with daclatasvir for the treatment of COVID-19. However, our study has a number of limitations; limitation of small sample size and the open-label rather than a blinded design, and limitation of infrequent PCR testing from nasal/pharyngeal swabs, which did not enable accurate measuring of time to virus negativity as we planned. We suggest that these results could be combined in a meta-analysis with other trials investigating sofosbuvir/daclatasvir and future trials of sofosbuvir/ daclatasvir in larger sample sizes should be conducted in early phase of infection (during the first week of infection). This could strengthen the evidence leading to regulatory approval of the use of SOF/DCV against COVID-19, for cases presenting early during the first week of illness.

## CONCLUSION

This study supports the potential benefits and safety of sofosbuvir combined with daclatasvir when given early in the treatment of COVID-19. We hope to motivate further large sized, multinational studies to confirm these benefits in order to add more options for the treatment of this serious infection that has been keeping the whole world in a dreadful crisis that all scientists should strive to put an end to.

## Data Availability

All data produced in the present study are available upon reasonable request to the authors.

## AUTHOR CONTRIBUTIONS

MY, SH conceived the idea, MY designed and wrote the protocol. BE, AZ, MF, ER, MH recruited the patients and collected the data. MY analyzed the data and prepared the first draft of the manuscript. All authors shared in interpreting the data, provided review and approved of the manuscript.

